# Global research trends and emerging fronts in refractory and macrolide-resistant *Mycoplasma pneumoniae* pneumonia in children: a bibliometric analysis (2000–2025)

**DOI:** 10.64898/2026.08.25.26361371

**Authors:** Deze Li, Hao Chen, Chen Shen

## Abstract

**Background:** Refractory and macrolide-resistant *Mycoplasma pneumoniae* pneumonia (MPP) has emerged as a major challenge in pediatric respiratory medicine, amplified by the post-2023 resurgence. However, a systematic overview of the research landscape specific to treatment-refractory and drug-resistant disease in children remains lacking.

**Methods:** Research articles and reviews on pediatric refractory or macrolide-resistant MPP published between 2000 and 2025 were retrieved from OpenAlex using Boolean searches. After screening, 2,286 records were quantitatively analyzed for annual output, contributing countries/institutions, thematic clusters, and citation-burst dynamics using Python.

**Results:** Annual publications grew exponentially, with a pronounced surge after 2023 (n=378 in 2025). China produced the highest volume (45.1%) but recorded fewer citations per publication than the US, Japan, and Canada. The literature resolved into four clusters: macrolide resistance/molecular basis, epidemiology, etiology/co-infection, and refractory disease management. Burst analysis showed an evolution from earlier fronts like 23S rRNA mutations and azithromycin to recent emerging trends like pandemic-related co-circulation, genotype surveillance, and co-infection.

**Conclusions:** Research on pediatric refractory and resistant MPP is expanding rapidly, shifting in emphasis from etiologic descriptions toward resistance mechanisms and clinical management. Standardizing the treatment of macrolide-unresponsive disease and post-pandemic epidemiological surveillance represent the principal directions for future work.

## Introduction

*Mycoplasma pneumoniae* (MP) is one of the most common causes of community-acquired pneumonia (CAP) in children and adolescents, accounting for a substantial proportion of hospitalized pediatric CAP, particularly among school-aged children[1, 2]. Although MP infection is often self-limiting, a clinically important subset of children develops refractory MPP — disease that fails to respond to appropriate macrolide therapy, with persistent fever, progressive radiographic consolidation, and an elevated risk of extrapulmonary complications and long-term sequelae such as bronchiolitis obliterans[3, 4].

Two developments have intensified clinical and scientific attention to this subset. First, macrolide resistance in MP, mediated predominantly by point mutations in the 23S rRNA gene, has reached high prevalence across East Asia and is rising in other regions, undermining first-line therapy and driving interest in alternative agents such as tetracyclines and fluoroquinolones, whose use in children requires careful risk–benefit assessment [5–7]. Second, following the relaxation of non-pharmaceutical interventions associated with the COVID-19 pandemic, many countries reported a marked resurgence of MP infection from 2023 onward, frequently accompanied by severe and treatment-refractory presentations[8, 9]. Together, these developments have produced a rapidly growing but fragmented body of literature.

Bibliometric analysis offers a quantitative, reproducible means of characterizing the structure and evolution of a research field, identifying influential contributors and works, and detecting emerging fronts. Previous bibliometric studies have examined MPP in children at a broad level, but to our knowledge no study has focused specifically on the refractory and drug-resistant subset, which constitutes the principal clinical and translational challenge[10, 11]. To address this gap, we conducted a bibliometric analysis of the literature on pediatric refractory and macrolide-resistant MPP from 2000 to 2025, aiming to (i) describe publication trends and overall productivity, (ii) identify leading countries, institutions, authors and journals, (iii) map the thematic structure through keyword co-occurrence clustering, and (iv) trace the evolution of research fronts through citation-burst analysis.

## Materials and methods

### Data source and search strategy

Bibliographic records were retrieved from OpenAlex, an open, comprehensive scholarly database that indexes works, authors, institutions and citations. The search was restricted to the title and abstract fields (‘title_and_abstract.search’) using the following Boolean strategy:

(*Mycoplasma Pneumoniae*) **AND** (child OR children OR pediatric OR paediatric OR infant OR adolescent) **AND** (refractory OR macrolide OR resistant OR resistance OR severe OR “drug resistance”)

This design combines a pathogen concept, a pediatric-population concept, and a refractory/resistance concept, so that the retrieved corpus is centred on the treatment-refractory and drug-resistant subset rather than MP infection in general.

### Inclusion and exclusion criteria

Records were limited to research articles and reviews published from 2000 onward. After retrieval, records were de-duplicated by DOI (with title-based de-duplication for records lacking a DOI), and entries without a title or publication year were removed. The screening flow is summarized in the Supplementary sample-overview table. A total of **2286** records met the criteria (2019 in the complete years 2000–2025; 267 in the partial year 2026, which was flagged separately and excluded from trend fitting). The corpus comprised 2117 research articles and 169 reviews.

### Analysis and visualization

All analyses were performed in Python. Bibliographic metadata were processed with pandas; ISO country codes were mapped to country names with pycountry; keyword co-occurrence networks were constructed and clustered with networkx, using the Louvain modularity algorithm for community detection; and figures were produced with matplotlib. Metrics included annual and cumulative publication counts, an exponential (log-linear) growth fit with a short-term forecast, total and mean citations, the h-index, the numbers of contributing countries and journals, per-entity productivity and citation impact, keyword co-occurrence clusters, and keyword burst strength based on year-on-year deviation of relative term frequency from each term’s overall baseline.

## Results

### Publication output and overall profile

The 2286 included records were cited 42,389 times in total (mean 18.5 citations per publication) and were distributed across 94 countries or regions and 712 journals, indicating a broad and internationally active field (Table 1).

**Table 1.** Overall bibliometric profile of the pediatric refractory/resistant MPP literature (2000–2025).

| Indicator | Value |
| --- | --- |
| Total publications (N) | 2286 |
| Publications in complete years (2000–2025) | 2019 |
| Publications in partial year (2026) | 267 |
| Research articles | 2117 |
| Reviews | 169 |
| Total citations | 42389 |
| Mean citations per publication | 18.5 |
| h-index | 94 |
| Contributing countries/regions | 94 |
| Journals | 712 |
| English-language (%) | 93.6 |

*Mycoplasma pneumoniae pneumonia. The h-index is computed over all included publications. Citation counts are cumulative as indexed by OpenAlex at the time of retrieval. The partial year 2026 is reported separately and excluded from trend analyses. Data source: OpenAlex.*

Annual output was low and relatively flat through the 2000s, rose steadily to 70 papers in 2015, and thereafter followed an approximately exponential trajectory. The most striking feature is the post-2023 surge: annual output roughly doubled in 2024 (n=189) and reached 378 in 2025 (Figure 1a), coinciding with the widespread resurgence of MP infection. A log-linear fit over 2010–2025 captured this exponential growth (Figure 1b) and projected continued high output through 2026–2028.

**Figure 1.**
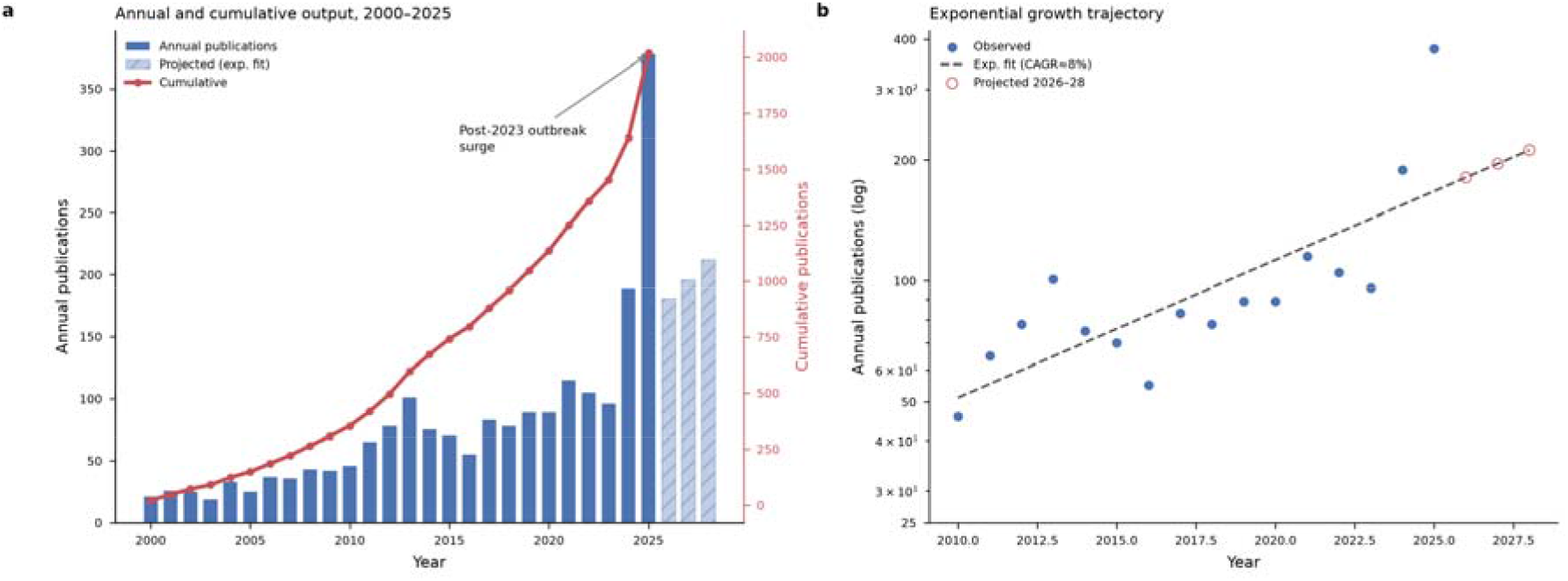
Annual and cumulative publication output on pediatric refractory and macrolide-resistant MPP, 2000–2025. **(a)** Annual publication counts (solid bars, left axis) and cumulative counts (red line, right axis); hatched bars show the projection for 2026–2028 from an exponential model fitted to 2010–2025 data; the arrow marks the surge following the 2023 resurgence of MP infection. **(b)** Annual counts on a logarithmic axis with the fitted exponential trajectory (log-linear least-squares fit, 2010–2025) and the 2026–2028 projection. Based on 2019 records published in the complete years 2000– 2025 (the partial year 2026 was excluded from trend fitting). Data source: OpenAlex.

### Contributions of countries, institutions and authors

China was by far the most productive country (1030 papers), followed by the United States (308) and South Korea (101) (Figure 2a; Table 2). At the institutional level, output was dominated by major Chinese pediatric centres, led by Beijing Children’s Hospital (84 papers), Capital Medical University (76) and Soochow University (53) (Figure 2b; Table 3). The most prolific authors included Zhimin Chen (23 papers, 1017 citations), Zhengrong Chen and Chao Yan, alongside internationally recognized contributors such as Patrick M. Meyer Sauteur (Figure 2c; Table 4).

**Figure 2.**
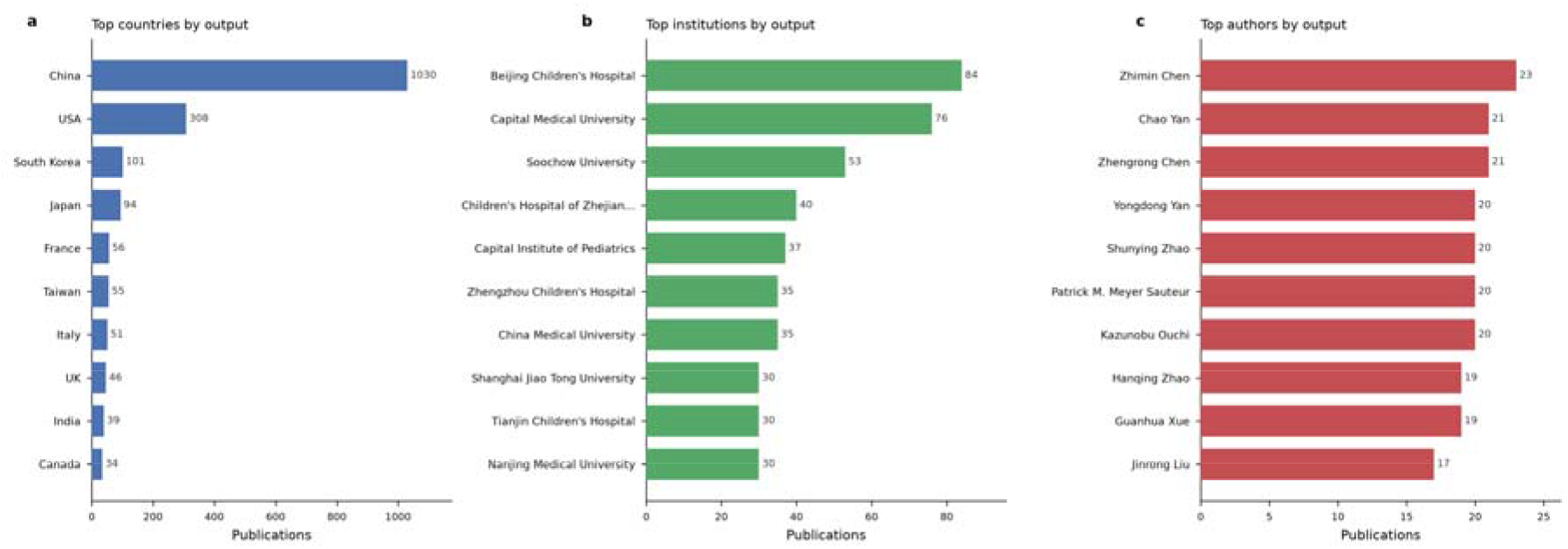
Leading contributors to the pediatric refractory/resistant MPP literature by publication count. **(a)** Top 10 countries, **(b)** top 10 institutions, and **(c)** top 10 authors. Bars show the number of publications; each country, institution, or author was counted once per publication (full counting). Values at the bar ends denote publication counts. Based on all 2286 included records. Data source: OpenAlex.

**Table 2.** Top 10 countries by publication output.

| Rank | Country/region | Publications | Citations | Citations/paper |
| --- | --- | --- | --- | --- |
| 1 | China | 1030 | 13611 | 13.2 |
| 2 | USA | 308 | 13163 | 42.7 |
| 3 | South Korea | 101 | 2748 | 27.2 |
| 4 | Japan | 94 | 3678 | 39.1 |
| 5 | France | 56 | 1870 | 33.4 |
| 6 | Taiwan | 55 | 1637 | 29.8 |
| 7 | Italy | 51 | 1563 | 30.6 |
| 8 | UK | 46 | 1774 | 38.6 |
| 9 | India | 39 | 294 | 7.5 |
| 10 | Canada | 34 | 2430 | 71.5 |

*Countries were assigned by author affiliation and counted once per publication (full counting); a publication with authors from multiple countries contributes to each. Citations/paper = total citations ÷ publications. Data source: OpenAlex.*

**Table 3.** Top 10 institutions by publication output.

| **Rank** | **Institution** | **Publications** | **Citations** | **Citations/paper** |
| --- | --- | --- | --- | --- |
| **1** | Beijing Children's Hospital | 84 | 1362 | 16.2 |
| **2** | Capital Medical University | 76 | 1720 | 22.6 |
| **3** | Soochow University | 53 | 984 | 18.6 |
| **4** | Children's Hospital of Zhejiang University | 40 | 1324 | 33.1 |
| **5** | Capital Institute of Pediatrics | 37 | 619 | 16.7 |
| **6** | Zhengzhou Children's Hospital | 35 | 323 | 9.2 |
| **7** | China Medical University | 35 | 512 | 14.6 |
| **8** | Shanghai Jiao Tong University | 30 | 567 | 18.9 |
| **9** | Tianjin Children's Hospital | 30 | 732 | 24.4 |
| **10** | Nanjing Medical University | 30 | 632 | 21.1 |

*Institutions were assigned by author affiliation as recorded in OpenAlex and counted once per publication. Institution names are reproduced as indexed and were not further disambiguated. Data source: OpenAlex.*

**Table 4.**
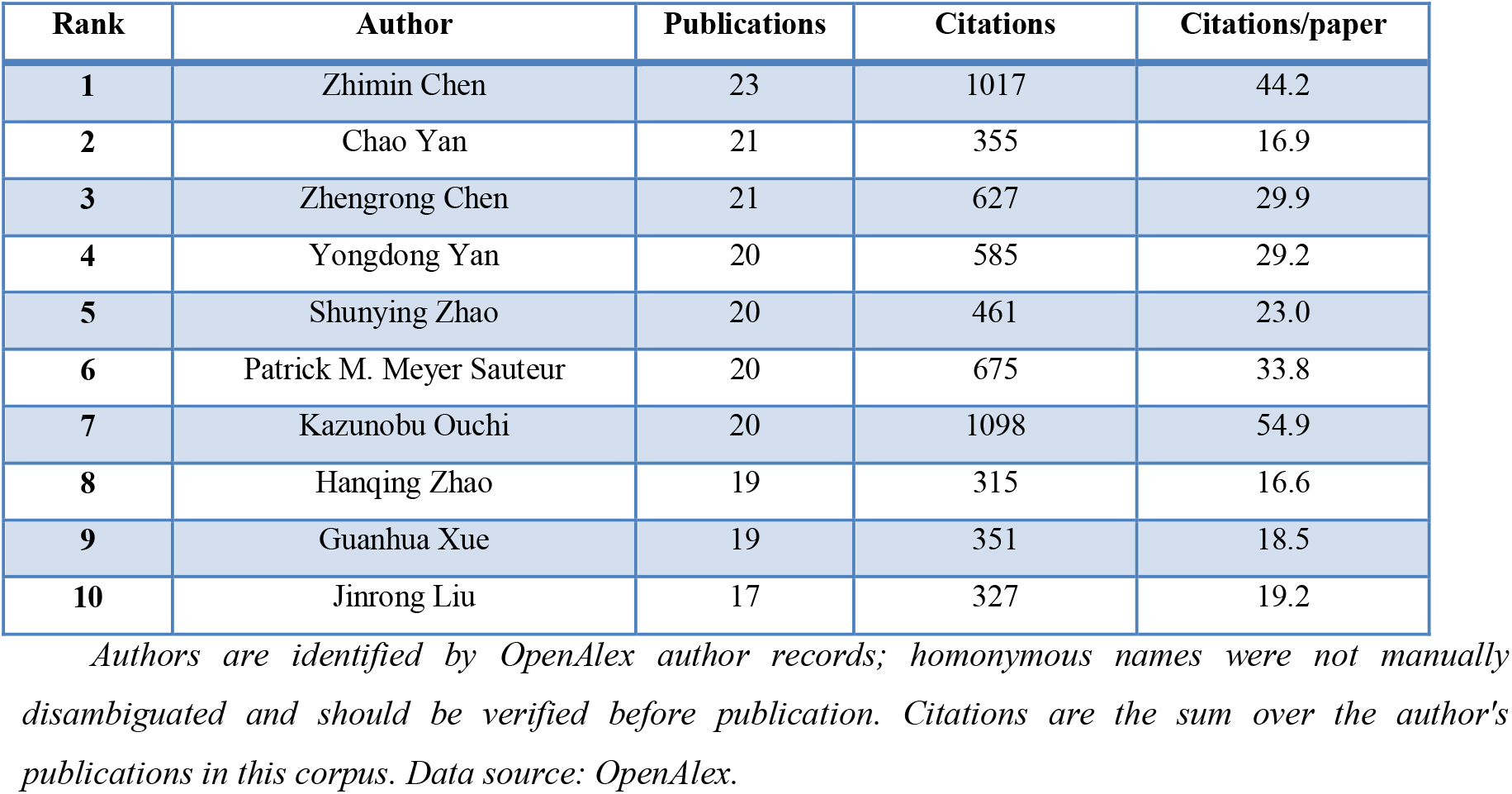
Top 10 most productive authors.

A comparison of output against citation impact revealed a clear divergence (Figure 3). Although China led output by a wide margin, its mean citations per publication (13.2) fell below the global average (18.5), whereas the United States (42.7), Japan and Canada achieved higher per-paper impact from smaller volumes. This pattern suggests that the rapid expansion of the field has been driven substantially by high-volume, often single-centre or retrospective, output, and highlights an opportunity to strengthen the international influence of the literature through higher-quality study designs.

**Figure 3.**
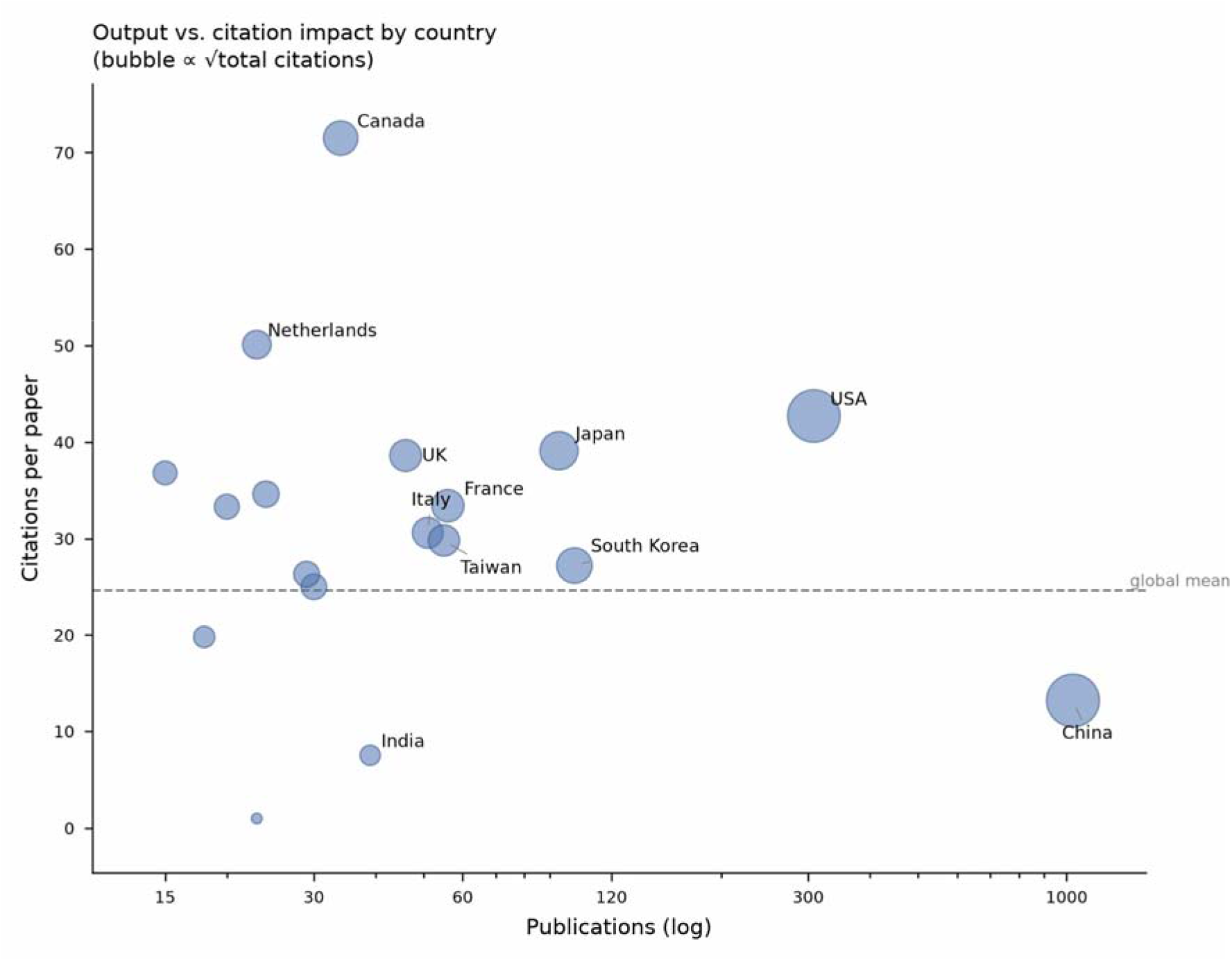
Publication output versus citation impact by country. The horizontal axis (logarithmic) shows the number of publications and the vertical axis shows the mean number of citations per publication; bubble area is proportional to the square root of a country’s total citations. The dashed line marks the global mean of 18.5 citations per publication. Only countries with ≥15 publications are shown. Citation counts are cumulative as indexed by OpenAlex at the time of retrieval. Data source: OpenAlex.

### Journals and highly cited papers

The most productive journals were pediatric and infectious-disease titles, led by *The Pediatric Infectious Disease Journal* (73 papers), *BMC Infectious Diseases* (64) and *Frontiers in Pediatrics* (63) (Figure 4a; Table 5). The most cited topic-relevant papers constitute the intellectual foundation of the field: authoritative reviews of MP as a human pathogen by Waites and colleagues, national and international guidelines for the management of community-acquired pneumonia, and landmark studies of macrolide resistance by Pereyre, Matsuoka, Kim and Morozumi (Figure 4b; Table 6)[12–14].

**Figure 4.**
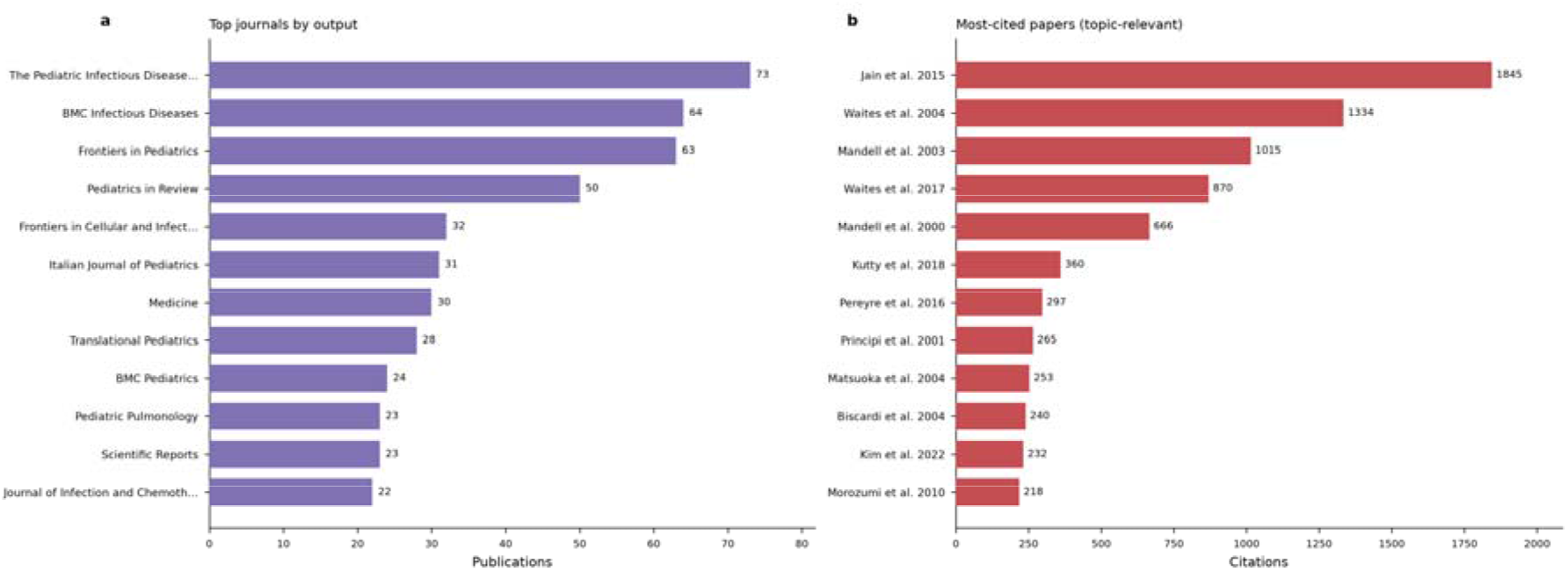
Most productive journals and most-cited papers. **(a)** Top 10 journals by number of publications. **(b)** The most-cited topic-relevant papers, labelled by first author and year. Topic-relevant papers were identified by title screening to exclude items retrieved by the search but unrelated to MP (e.g. COVID-19 treatment protocols); placeholder source records were removed. Citation counts are cumulative as indexed by OpenAlex at the time of retrieval. Data source: OpenAlex.

**Table 5.** Top 10 journals by publication output.

| Rank | Journal | Publications | Citations | Citations/paper |
| --- | --- | --- | --- | --- |
| 1 | The Pediatric Infectious Disease Journal | 73 | 2141 | 29.3 |
| 2 | BMC Infectious Diseases | 64 | 1722 | 26.9 |
| 3 | Frontiers in Pediatrics | 63 | 608 | 9.7 |
| 4 | Pediatrics in Review | 50 | 262 | 5.2 |
| 5 | Frontiers in Cellular and Infection Microbiology | 32 | 441 | 13.8 |
| 6 | The Italian Journal of Pediatrics/Italian journal of pediatrics | 31 | 407 | 13.1 |
| 7 | Medicine | 30 | 574 | 19.1 |
| 8 | Translational Pediatrics | 28 | 155 | 5.5 |
| 9 | BMC Pediatrics | 24 | 559 | 23.3 |
| 10 | Pediatric Pulmonology | 23 | 747 | 32.5 |

*Journals are the primary publication venue recorded in OpenAlex; placeholder source records were removed. Data source: OpenAlex.*

**Table 6.**
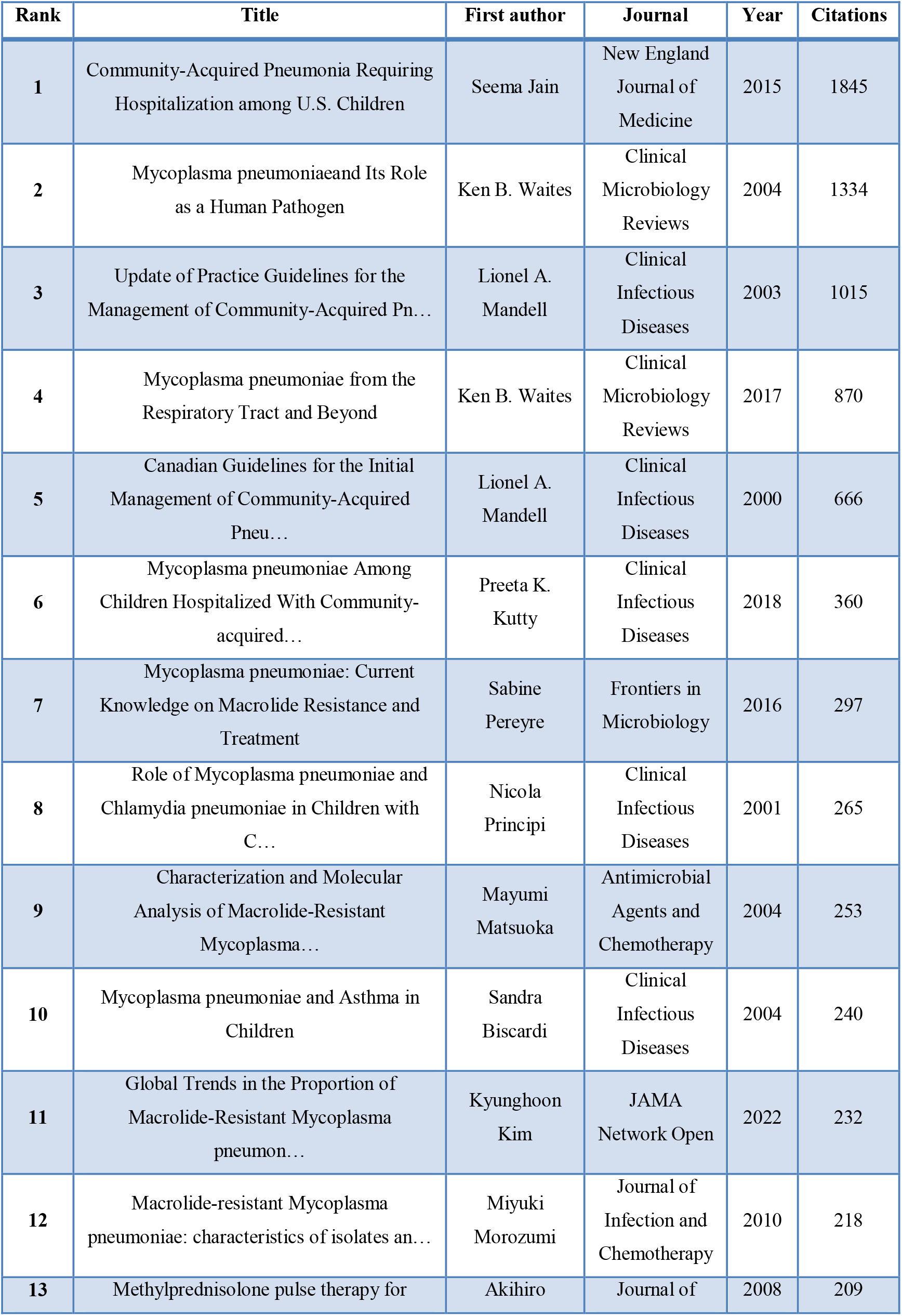
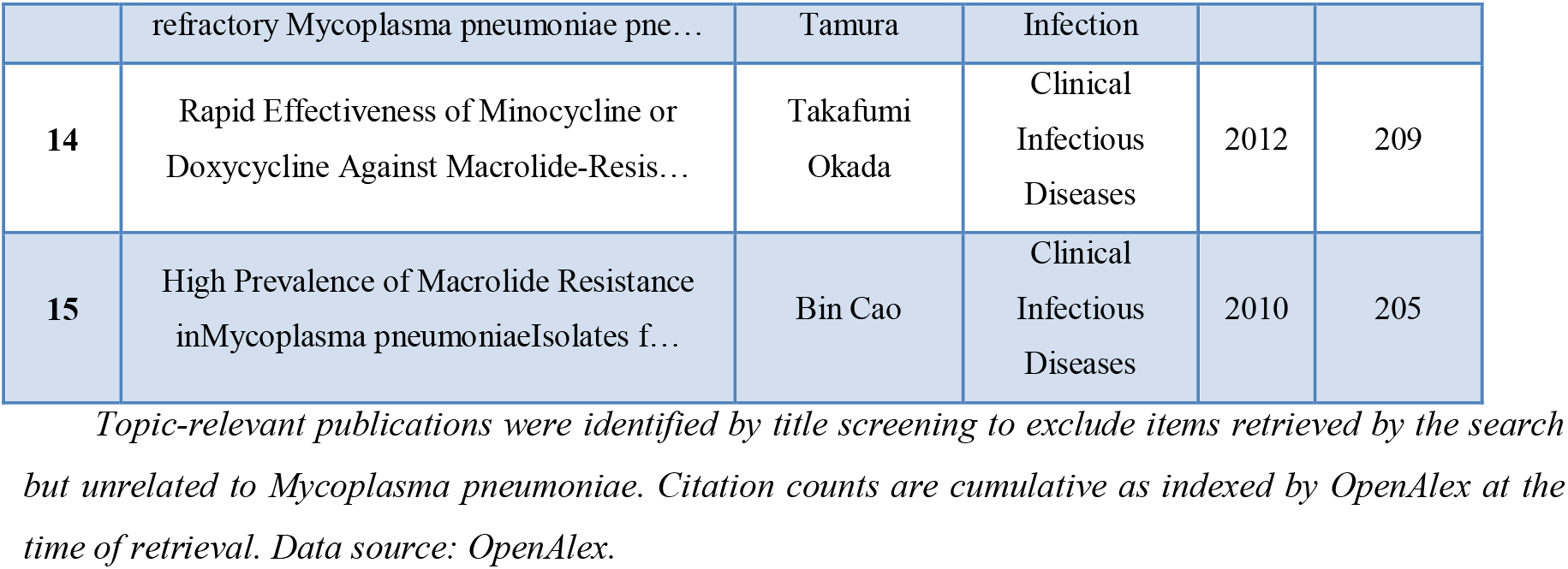
The 15 most-cited topic-relevant publications.

**Table 7.**
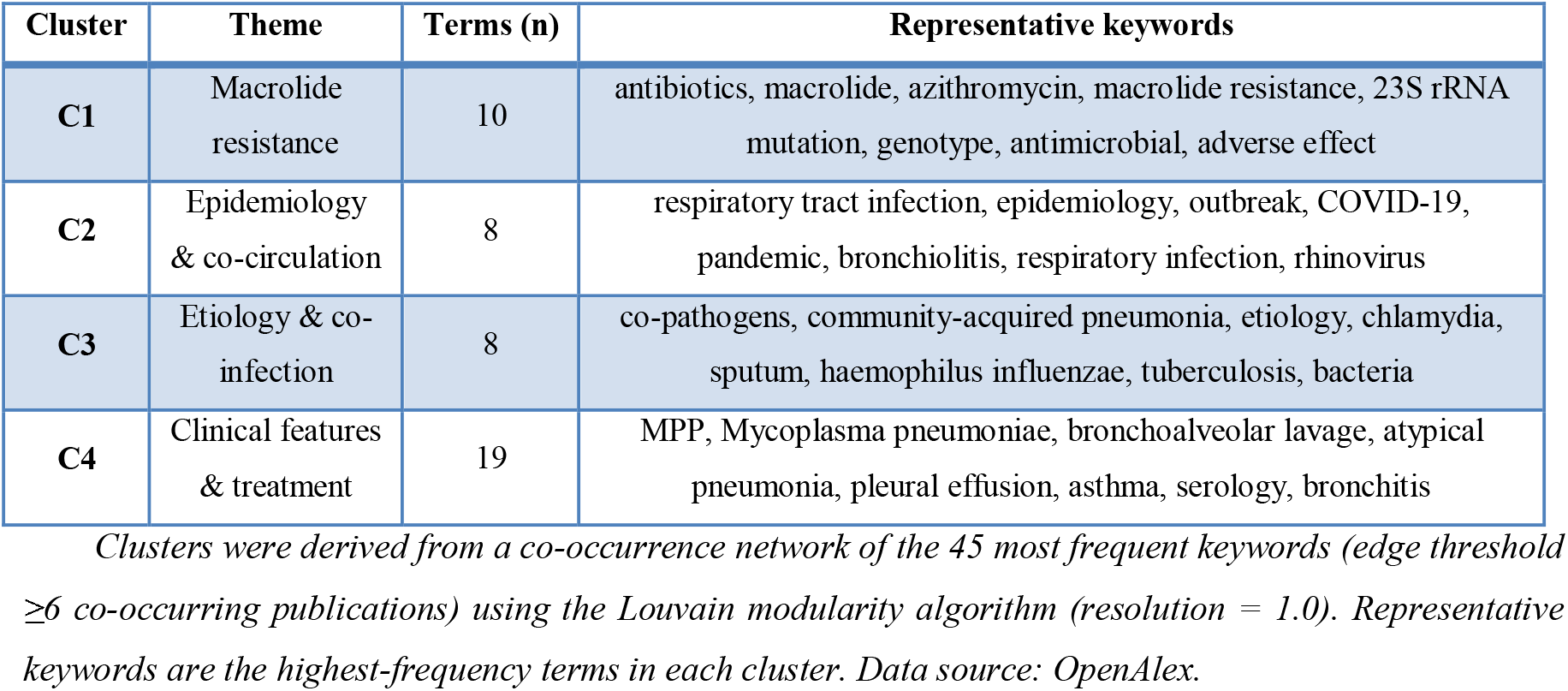
Thematic clusters identified by keyword co-occurrence analysis.

| Cluster | Theme | Terms (n) | Representative keywords |
| --- | --- | --- | --- |
| C1 | Macrolide resistance | 10 | antibiotics, macrolide, azithromycin, macrolide resistance, 23S rRNA mutation, genotype, antimicrobial, adverse effect |
| C2 | Epidemiology & co-circulation | 8 | respiratory tract infection, epidemiology, outbreak, COVID-19, pandemic, bronchiolitis, respiratory infection, rhinovirus |
| C3 | Etiology & co-infection | 8 | co-pathogens, community-acquired pneumonia, etiology, chlamydia, sputum, haemophilus influenzae, tuberculosis, bacteria |
| C4 | Clinical features & treatment | 19 | MPP, Mycoplasma pneumoniae, bronchoalveolar lavage, atypical pneumonia, pleural effusion, asthma, serology, bronchitis |

*Clusters were derived from a co-occurrence network of the 45 most frequent keywords (edge threshold $\geq 6$ co-occurring publications) using the Louvain modularity algorithm (resolution = 1.0). Representative keywords are the highest-frequency terms in each cluster. Data source: OpenAlex.*

The prominence of resistance-focused works among the most cited papers underscores the centrality of drug resistance to the field’s development.

### Thematic structure: keyword co-occurrence and clustering

Co-occurrence analysis of the 45 most frequent keywords yielded a network that resolved, by Louvain community detection, into four coherent thematic clusters (Figure 5):

1. **Macrolide resistance and its molecular basis** — antibiotics, macrolide, azithromycin, macrolide resistance, 23S rRNA mutation, genotype and alternative agents such as doxycycline.
2. **Epidemiology and pathogen co-circulation** — respiratory tract infection, epidemiology, outbreak, pandemic, COVID-19 and co-circulating pathogens.
3. **Etiology and co-infection** — community-acquired pneumonia, co-pathogens, etiology, chlamydia and *Haemophilus influenzae*.
4. **Clinical features and treatment** — refractory MPP, atypical pneumonia, pleural effusion, corticosteroid, bronchoalveolar lavage, serology and asthma.

**Figure 5.**
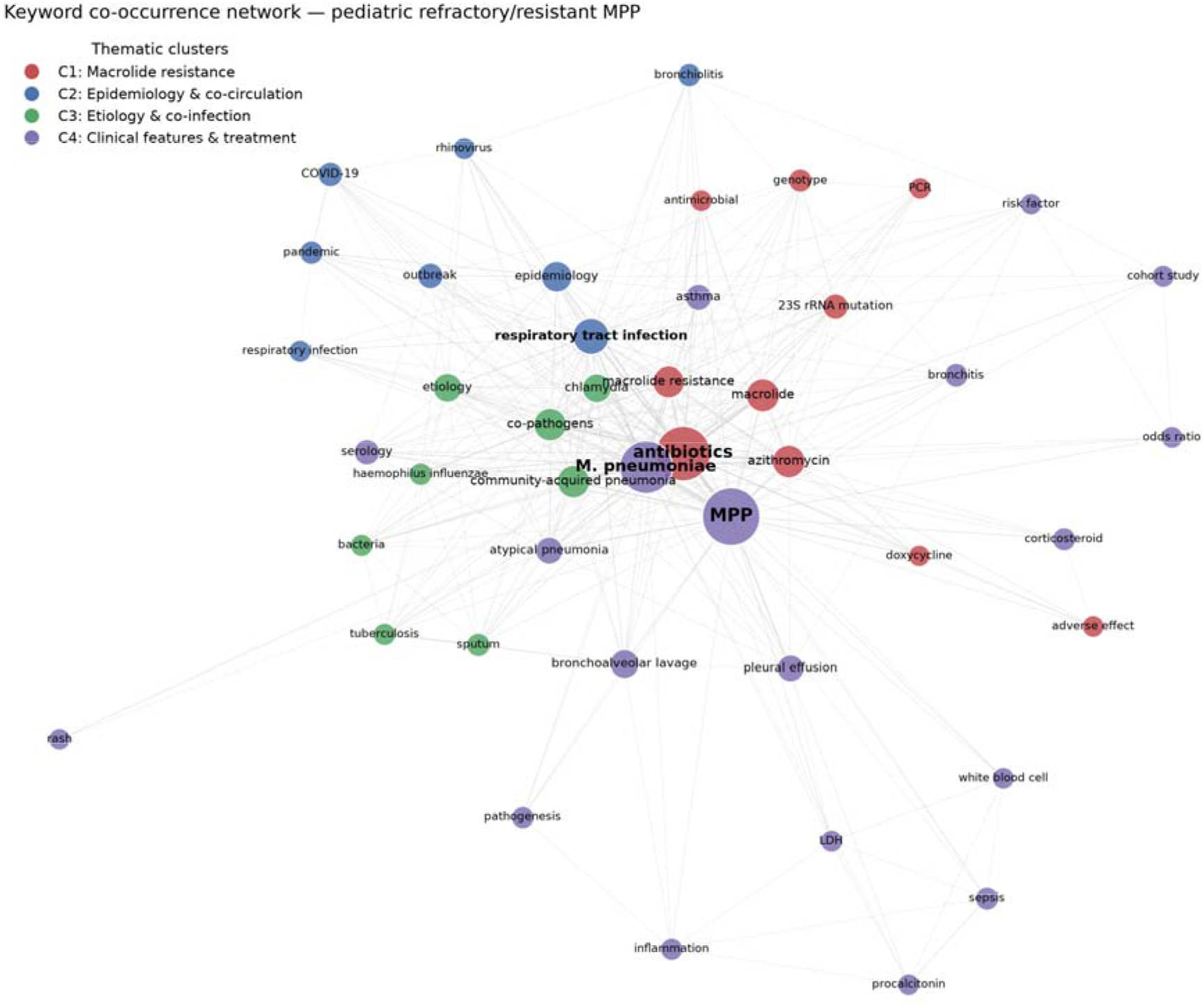
Keyword co-occurrence network. The network comprises the 45 most frequent author keywords and OpenAlex concepts after normalization and stop-word removal; edges connect terms co-occurring in ≥6 publications. Node size is proportional to term frequency and node colour indicates the thematic cluster identified by the Louvain modularity algorithm (resolution = 1.0): C1, macrolide resistance; C2, epidemiology and co-circulation; C3, etiology and co-infection; C4, clinical features and treatment. Based on all 2286 included records. Data source: OpenAlex.

These clusters map directly onto the principal axes of clinical and scientific concern: why disease becomes resistant, how it spreads, what co-infections accompany it, and how refractory presentations are recognized and managed.

### Research fronts: burst detection and thematic evolution

Burst analysis traced a clear progression of research fronts (Figure 6). Early fronts (mid-2000s to early 2010s) centred on etiology, serology and co-pathogens; a middle phase emphasized macrolide resistance, 23S rRNA mutation and azithromycin; and the most recent fronts (2022–2025) comprise pandemic- and COVID-19-related co-circulation, genotype surveillance, co-infection and bronchoalveolar lavage. The recency and strength of the pandemic-related terms reflect the field’s rapid reorientation toward the post-2023 epidemiological context.

**Figure 6.**
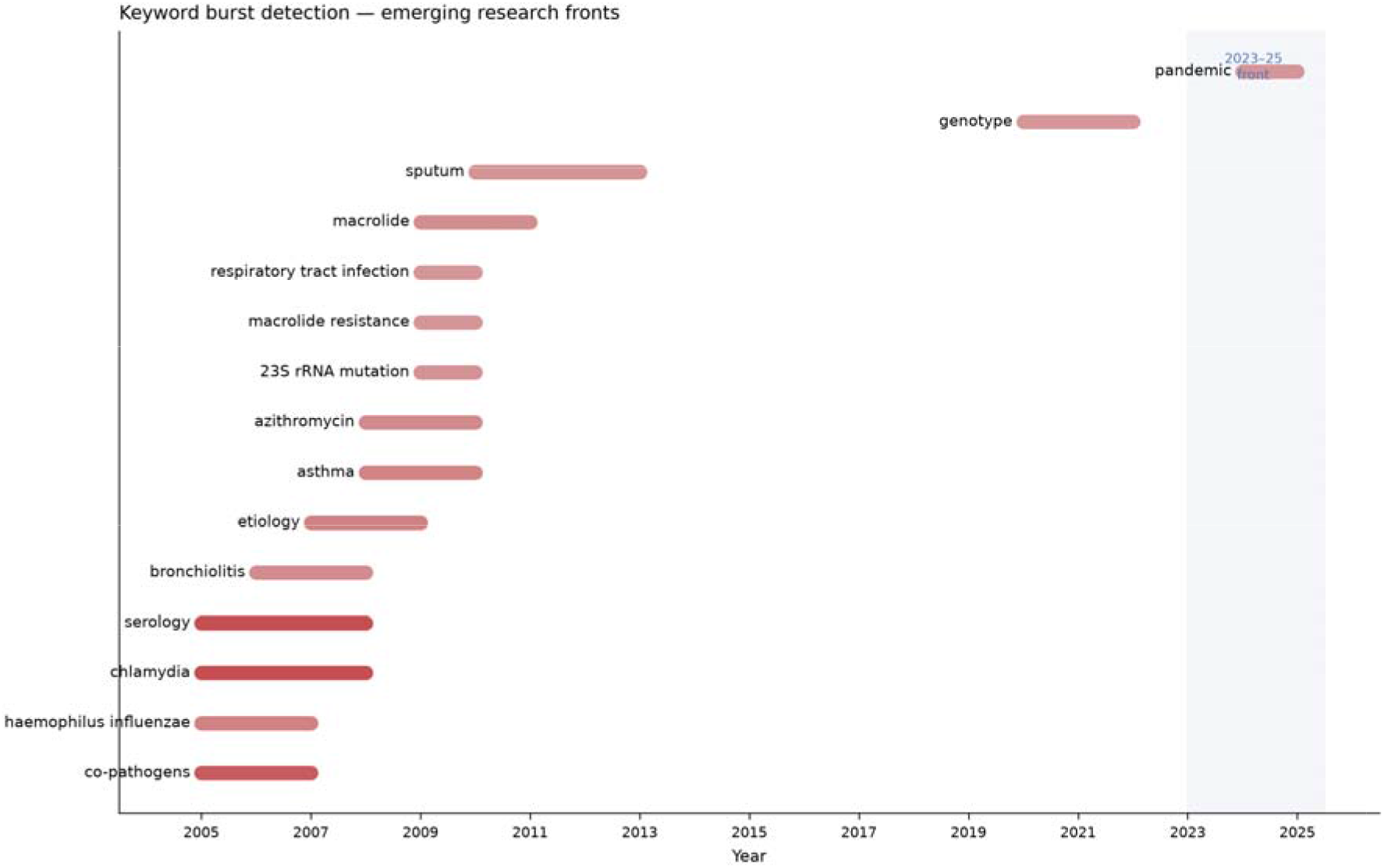
Keyword burst detection identifying emerging research fronts. Each horizontal bar spans the burst period of a term — the contiguous interval during which its yearly share of the themed literature most exceeded its overall baseline share (2005– 2025). Bar shade is proportional to burst strength (darker = stronger). Only terms occurring in ≥25 publications are considered; the omnipresent core terms (*Mycoplasma pneumoniae*, MPP, antibiotics) and non-substantive methodological terms were excluded. Data source: OpenAlex.

The shift in thematic emphasis is quantified in the thematic-evolution analysis (Figure 7). The share of keywords attributable to clinical features and treatment rose from roughly one-quarter of the themed literature in 2005 to close to half by 2024, becoming the dominant theme, while the etiology/co-infection share declined correspondingly. Macrolide resistance maintained a substantial and stable presence throughout, and epidemiology/co-circulation expanded in the most recent years.

**Figure 7.**
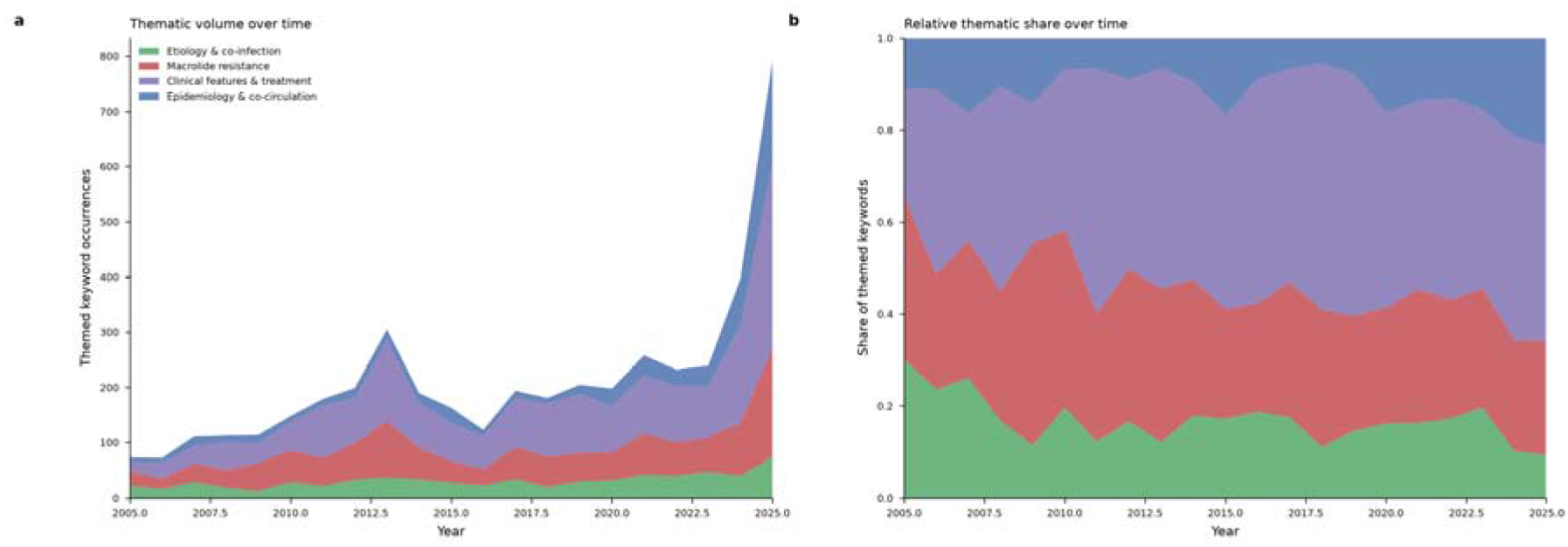
Evolution of research themes, 2005–2025. **(a)** Absolute number of themed-keyword occurrences per year by thematic cluster (stacked). **(b)** Relative share of each thematic cluster per year (stacked to 100%). Themes correspond to the four Louvain clusters in Figure 5. A themed keyword is any keyword assigned to one of the four clusters; the current partial year (2026) is excluded. Data source: OpenAlex.

## Discussion

This bibliometric analysis of 2286 publications provides the first field-level overview focused specifically on refractory and macrolide-resistant MPP in children. Four principal findings emerge.

Publication output has grown approximately exponentially since 2010 and surged sharply after 2023. The timing of this surge, concurrent with the widespread post-pandemic resurgence of MP infection and reports of severe and refractory disease, indicates that research activity is closely coupled to clinical burden. The projected continuation of high output through 2026–2028 suggests the topic will remain active in the near term.

China’s dominant share of publications, coupled with a below-average citation rate per paper, points to a literature built substantially on high-volume retrospective and single-centre reports. Strengthening multicentre collaboration and prospective, methodologically rigorous designs would be expected to raise the international influence and clinical translatability of this work. Notably, the highest-impact contributions in the field — including the global resistance-trend analysis of Kim et al. and the foundational pathogen reviews of Waites et al. combine large-scale synthesis with international authorship, in contrast to the predominantly single-centre Chinese output that drives publication volume[15, 16].

With a shift from description toward mechanism and management, the thematic-evolution analysis shows the centre of gravity moving from etiologic description toward resistance mechanisms and, increasingly, the clinical management of refractory disease. This reflects unmet clinical needs: how to treat children whose infection does not respond to macrolides, when to escalate to alternative antibiotics despite pediatric safety considerations, and how best to deploy adjunctive measures such as corticosteroids and bronchoscopic lavage[17, 18]. The emergence of a Chinese national expert consensus on macrolide-resistant MPP in 2024 and the proliferation of predictive nomograms for refractory disease signal a field-wide move from describing resistance toward operationalizing its management at the bedside and a transition also reflected in the rising share of clinical-treatment keywords documented here[19–21].

Molecular surveillance of 23S rRNA mutations and resistance genotypes, and post-pandemic epidemiological monitoring of MP co-circulation with other respiratory pathogens, are the most prominent recent fronts and are likely priorities for future research[3, 22, 23]. The concurrent 2023–2024 resurgence reported across Asia and Europe suggests these fronts are converging: understanding why the post-pandemic epidemic was accompanied by frequent macrolide-refractory presentations will require linking genotype surveillance to clinical outcomes, an integration that the current literature — fragmented across resistance, epidemiology, and clinical clusters — has yet to achieve[9, 24].

## Limitations

Several limitations should be considered. First, the analysis relied on a single database (OpenAlex); although its coverage is broad, results may differ modestly from those based on Web of Science or Scopus. Second, the search was based on title and abstract keywords, which may omit some relevant records or admit a small number of tangential ones; we mitigated the latter by applying topic-relevance filtering to the highly cited list and by curating the keyword set. Third, OpenAlex concept tags are algorithmically assigned and noisy; despite normalization and stop-word removal, some subjectivity remains in the cluster boundaries. Fourth, data for 2026 were incomplete and were excluded from trend fitting, and the growth projection is an extrapolation that should be interpreted cautiously. Finally, bibliometric indicators reflect research attention and citation behaviour rather than clinical evidence quality, and should not be read as measures of clinical validity.

## Conclusions

Over the past 25 years, research on refractory and macrolide-resistant MPP in children has grown rapidly, entering a new phase of intense activity following the post-2023 resurgence of infection. The field’s emphasis has shifted from etiologic description toward resistance mechanisms and the clinical management of refractory disease. Molecular surveillance of macrolide resistance, the standardization of treatment for macrolide-unresponsive disease, and post-pandemic epidemiological monitoring stand out as the principal directions for future research.

## Data Availability

The authors in the article/Supplementary Material will provide the original data that
support this paper's conclusions. Further inquiries can be directed to the corresponding
authors.

## Reference

1. Rueda ZV, Aguilar Y, Maya MA, López L, Restrepo A, Garcés C, Morales O, Roya-Pabón C, Trujillo M, Arango C, et al: Etiology and the challenge of diagnostic testing of community-acquired pneumonia in children and adolescents. 2022, 22:169.

2. Li D, Zheng H, Wang X, Li F, Wang H, Chen H, Shen C, research SZJI: Investigation of T lymphocyte subsets in children with Mycoplasma pneumoniae pneumonia. 2024, 73:24.

3. Li W, Gao W, Xiong X, Tang X, one ALJP: Association analysis of Mycoplasma pneumoniae 23S rRNA gene mutation with refractory Mycoplasma pneumoniae pneumonia in children. 2026, 21:e0341580.

4. Zhu L, Xu J, Yuan T, Li H, pediatrics JLJFi: Screening of early predictive serum biomarkers and construction of a combined predictive model for refractory Mycoplasma pneumoniae infection in children. 2026, 14:1680913.

5. Mandell LA, Marrie TJ, Grossman RF, Chow AW, America RHHJCidaopotIDSo: Canadian guidelines for the initial management of community-acquired pneumonia: an evidence-based update by the Canadian Infectious Diseases Society and the Canadian Thoracic Society. The Canadian Community-Acquired Pneumonia Working Group. 2000, 31:383–421.

6. Matsuoka M, Narita M, Okazaki N, Ohya H, Yamazaki T, Ouchi K, Suzuki I, Andoh T, Kenri T, Sasaki Y, et al: Characterization and molecular analysis of macrolide-resistant Mycoplasma pneumoniae clinical isolates obtained in Japan. 2004, 48:4624–4630.

7. science KWYJJoKm: Growing Threat of Macrolide-Resistant Mycoplasma pneumoniae Among Children: What We Know and What We Need. 2025, 40:e317.

8. Liu R, Shao W, Qing Ye %J APMIS : acta pathologica m, et immunologica Scandinavica: Resurgence of Mycoplasma pneumoniae Infections in the Post-COVID-19 Era: Epidemiology, Therapeutic Challenges, and Mitigation Strategies. 2025, 133:e70092.

9. Microbe EMsgJTL: Global spatiotemporal dynamics of Mycoplasma pneumoniae re-emergence after COVID-19 pandemic restrictions: an epidemiological and transmission modelling study. 2025, 6:101019.

10. Pereyre S, Goret J, microbiology CBJFi: Mycoplasma pneumoniae: Current Knowledge on Macrolide Resistance and Treatment. 2016, 7:974.

11. Zhou Y, Wang J, Chen W, Shen N, Tao Y, Zhao R, Luo L, Li B, diseases QCJBi: Impact of viral coinfection and macrolide-resistant mycoplasma infection in children with refractory Mycoplasma pneumoniae pneumonia. 2020, 20:633.

12. Waites KB, reviews DFTJCm: Mycoplasma pneumoniae and its role as a human pathogen. 2004, 17:697-728, table of contents.

13. Cao B, Zhao C-J, Yin Y-D, Zhao F, Song S-F, Bai L, Zhang J-Z, Liu Y-M, Zhang Y-Y, Wang H, America CWJCidaopotIDSo: High prevalence of macrolide resistance in Mycoplasma pneumoniae isolates from adult and adolescent patients with respiratory tract infection in China. 2010, 51:189–194.

14. Morozumi M, Takahashi T, infection KUJJo, Chemotherapy cojotJSo: Macrolide-resistant Mycoplasma pneumoniae: characteristics of isolates and clinical aspects of community-acquired pneumonia. 2010, 16:78–86.

15. Kim K, Jung S, Kim M, Park S, Yang H-J, open ELJJn: Global Trends in the Proportion of Macrolide-Resistant Mycoplasma pneumoniae Infections: A Systematic Review and Meta-analysis. 2022, 5:e2220949.

16. Waites KB, Xiao L, Liu Y, Balish MF, reviews TPAJCm: Mycoplasma pneumoniae from the Respiratory Tract and Beyond. 2017, 30:747–809.

17. Ding G, Zhang X, Vinturache A, Rossum AMCv, Yin Y, pediatrics YZJEjo: Challenges in the treatment of pediatric Mycoplasma pneumoniae pneumonia. 2024, 183:3001–3011.

18. Blyth CC, Society JSGJJotPID: Macrolides in Children With Community-Acquired Pneumonia: Panacea or Placebo? 2018, 7:71–77.

19. Choi YJ, Chung EH, Lee E, Kim C-H, Lee YJ, Kim H-B, Kim B-S, Kim HY, Cho Y, Seo J-H, et al: Clinical Characteristics of Macrolide-Refractory Mycoplasma pneumoniae Pneumonia in Korean Children: A Multicenter Retrospective Study. 2022, 11.

20. Huang W, Xu X, Zhao W, research QCJJoi: Refractory Mycoplasma Pneumonia in Children: A Systematic Review and Meta-analysis of Laboratory Features and Predictors. 2022, 2022:9227838.

21. Wang Y-S, Zhou Y-L, Bai G-N, Li S-X, Xu D, Chen L-N, Chen X, Dong X-Y, Fu H-M, Fu Z, et al: Expert consensus on the diagnosis and treatment of macrolide-resistant Mycoplasma pneumoniae pneumonia in children. 2024, 20:901–914.

22. Centrone F, Accogli M, Melilli R, Marziani A, Orlando VA, Casulli D, Scolamacchia V, Netti N, Sacco D, Sallustio A, et al: Prevalence of Macrolide-Resistant Mycoplasma pneumoniae Infections After the COVID-19 Pandemic in Southern Italy, 2023-2025. 2025, 14:2733–2741.

23. Xu M, Li Y, Shi Y, Liu H, Tong X, Ma L, Gao J, Du Q, Du H, Liu D, et al: Molecular epidemiology of Mycoplasma pneumoniae pneumonia in children, Wuhan, 2020-2022. 2024, 24:23.

24. Yan C, Tong S, Wu Y, Chen Y, Jia X, Guo Y, Cui M, Pei G, Zhang Z, Zhou H, et al: Macrolide-resistant Mycoplasma pneumoniae resurgence in Chinese children in 2023: a longitudinal, cross-sectional, genomic epidemiology study. 2025, 6:101200.

